# Integrating polygenic and transcriptional risk scores improves risk prediction of nine common diseases in the underrepresented Vietnamese population

**DOI:** 10.64898/2025.12.08.25341869

**Authors:** Sang V. Nguyen, Tien M. Pham, Tham H. Hoang, Trang T. H. Tran, Giang M. Vu, Mai H. Tran, Thien K. Nguyen, Huy Le Trinh, Huyen Thi Thanh Vu, Tuan Minh Pham, Dung Trung Nghiem, Anh Gia Pham, Yen Hoang, Giang H. Phan, Dat X. Dao, Hung N. Luu, Tran Huy Thinh, Quan Nguyen, Buu Truong, Nam S. Vo

**Affiliations:** Center for Biomedical Informatics, Vingroup Big Data Institute, Hanoi, Vietnam; R&D Department, GeneStory JSC, Hanoi, Vietnam; Department of Oncology, Hanoi Medical University, Hanoi, Vietnam; Faculty of Geriatrics, Hanoi Medical University, Hanoi, Vietnam; National Geriatric Hospital, Hanoi, Vietnam; Department of Cardiology, Hanoi Medical University, Hanoi, Vietnam; Vietnam National Heart Institute; Bach Mai Hospital, Hanoi, Vietnam; Viet-Duc Hospital, Hanoi, Vietnam; Department of Science and Technology Management, Hanoi Medical University, Hanoi, Vietnam; Department of Biochemistry, Hanoi Medical University; UPMC Hillman Cancer Center, University of Pittsburgh Medical Center, Pittsburgh, PA 15232, USA; Department of Epidemiology, School of Public Health, University of Pittsburgh, Pittsburgh, PA 15261, USA; Dr. Mary and Ron Neal Cancer Center, Houston Methodist Research Institute, Houston Methodist Hospital, Houston, TX 77030, USA; QIMR Berghofer Medical Research Institute and University of Queensland, Brisbane, QLD 4072, Australia; Department of Epidemiology, Harvard T.H. Chan School of Public Health, Boston, MA, 02115; Program in Medical and Population Genetics, Broad Institute of MIT and Harvard, Cambridge, MA, 02142; VinUni Big Data Research Institute, VinUniversity, Hanoi, Vietnam

## Abstract

Polygenic risk scores (PRS) represent the cumulative impact of numerous common genomic variants, to predict clinical phenotypes and outcomes for individuals. However, PRS are typically derived from GWAS for populations of European origin, often resulting in reduced performance and their transferability to other underserved populations. In this study, we comprehensively analyzed 550 samples for nine common diseases in the Vietnamese population, including breast cancer (BC), colorectal cancer (CRC), gastric cancer (GC), chronic kidney disease (CKD), coronary artery disease (CAD), hyperlipidemia, osteoporosis, osteoarthritis, and Parkinson’s disease (PD). Healthy control subjects were taken from the 1000 Vietnamese Genomes Project (VN1K). We evaluated seven advanced PRS algorithms using multiple GWAS datasets from both East Asian and European populations and identified the best performing method for each disease. PRS accuracy, assessed by incremental liability R-squared (incR2), ranged from 1.8% in CKD to 8.3% in CAD. The Area Under the Curve (AUC) ranged from 0.55 for CKD to 0.70 for CAD. Integrating with transcriptional risk scores (TRS), the PRS+TRS model led to a consistently increased incR2 across all nine diseases ranging from 1% to 15%. These findings offer valuable insights into the implementation of PRS+TRS for disease risk prediction in the Vietnamese population, where a similar approach would be applicable to other underrepresented populations.

## INTRODUCTION

Biomedical research has been utilizing the large genomic information to understand how variation in genotypes may lead to disease phenotypes. The genome-wide association studies (GWAS) have proven the potential to discover important genomic markers ^1, 2^. However, most GWAS have focused on populations with European origin (>78% in the UK biobank and >85% in the GTEx project) ^3, 4^. Therefore, this ancestral disproportion in GWAS raises growing concerns about inequities in global genomic medicine. Many of the common alleles displayed in the global reference genome (e.g., GRCh38) were determined based on populations of European descendants. These common alleles can be rare alleles in underrepresented populations, leading to mis-calling or false positive variants ^5^. Furthermore, many existing models and bioinformatics tools, particularly those for cardiometabolic and other common complex diseases, are predominantly derived from European cohorts, which may introduce biases when applied to populations with different genetic backgrounds ^6, 7^.

Polygenic risk scores (PRS) quantify an individual’s genomic predisposition to complex traits and diseases by combining effect sizes of genomic variants in the traits learnt from GWAS. These scores are based on the cumulative effects of multiple genomic variants, each contributing a small amount to the overall risk; thus, PRS offers an approach to integrate information from GWAS that can be used in the construction of risk predictive models ^8, 9, 10^. By evaluating the presence of specific single nucleotide polymorphisms (SNPs) and their associated effect sizes, PRS helps to identify individuals who are at higher or lower risk for conditions such as cardiovascular diseases, metabolic diseases, and cancers relative to a specific population ^6, 11, 12^. Complementing PRS, transcriptional risk scores (TRS) incorporate gene expression data by linking genomic variants from GWAS to transcriptional activity through expression quantitative trait loci (eQTL) through colocalization method ^13^. This integration adds a functional layer to genomic risk, enhancing the predictive power of PRS by accounting for downstream molecular effects of genomic variants ^14,15^. Together, combining both PRS and TRS deepens our understanding of the genomic architecture underlying diseases and supports the advancement of personalized medicine by enabling more targeted prevention and treatment strategies based on an individuals’ unique genomic profile.

Although PRS and TRS have been extensively studied for common diseases globally, systematic research focused on the Vietnamese population remains limited. As PRS research advances, there is growing recognition of the need to validate and refine these models across diverse ancestral backgrounds ^16^. Emerging studies have shown that utilizing GWAS data from populations closely related to the target population (which is less studied) and European populations, combined with advanced PRS calculation techniques, can significantly improve predictive accuracy in under-studied populations ^17, 18^. These studies have also highlighted the importance of customizing bioinformatics pipelines and statistical approaches to account for unique linkage disequilibrium patterns and allele frequency distributions in non-European populations. Our work was based on the rationale that by integrating multi-ethnic GWAS, state-of-the-art PRS and TRS methodologies, and population-specific disease datasets, researchers can better characterize disease risk for individuals and potentially uncover novel risk factors that are specific to these under-studied populations.

Addressing this gap is crucial and the transethnic transferability of PRS remains a major challenge. Therefore, systematic evaluations of this approach for underrepresented populations, such as the Vietnamese, are essential to ensure equitable access to genomics-driven personalized medicine. The 1000 Vietnamese Genomes Project (VN1K) plays a pioneering role in reducing the inequality in genomic data research and applications ^19^. This database allows researchers to identify these variants that are specific for Vietnamese and have not been reported in previous studies which were biased towards European descendants. Furthermore, VN1K provides a foundation for developing and evaluating risk prediction models tailored to improve disease prediction in this underrepresented group.

In this study, we leveraged GWAS data from East Asia (EAS) and Europe (EUR) to analyze genotyping data of 550 samples collected from patients of nine common diseases in Vietnam and whole genome sequencing data of 1008 healthy individuals from VN1K. We developed a comprehensive workflow to process data, calculate PRS and perform benchmarking for each of the diseases. Our pipeline used the most advanced methods, available to date, including PRSice-2 ^20^, LDpred2 ^21^, PRS-CS ^22^, PRS-CSx ^23^ and shaPRS ^24^; and their combinations, including shaPRS+LDpred2 ^21, 24^ and shaPRS+ PRS-CSx ^23, 24^. Additionally, we curated SNP effect size datasets from the PGS Catalog for each disease to calculate PRS and compared them with PRS models from different tested methods. To evaluate the performance of such PRS models for binary phenotypes, we used liability scale-adjusted pseudo-R² to facilitate comparisons, along with AUC (Area Under the Curve), a common and well-established metric for binary classification assessment ^25^. We included GWAS data from populations that were closely related to the Vietnamese, including BioBank Japan (BBJ) ^26^ and other East Asian populations, as well as cross-ethnic studies of European populations. We also leveraged transcriptional data, such as aggregating gene expression levels linked to risk alleles by utilizing tissue-specific eQTL data from the GTEx study ^27^, to improve the accuracy of the risk prediction models. Considering possible confounding factors, we thoroughly examined population stratification of disease risk across different age groups and genders. Taken together, our study sought to offer comprehensive insights into the application of PRS and TRS methodologies using a rich dataset on diseases and a population that were genomically under-studied (such as the Vietnamese population). We expect that our work will be applicable to other populations with under-represented genomics data such as those in Asia, Latin America and Africa.

## RESULTS

### Overview of the studied cohorts

We first examined the baseline demographics of the case cohorts for the nine investigated diseases and the control cohort (VN1K, N=1,008) (**Table 1**). The total number of case samples included in these analyses was 550. On the one hand, the case cohorts of the nine diseases were older individuals compared to the VN1K controls. For example, the mean age was notably higher in disease groups such as coronary artery disease (N=50, Mean=66.7, SD=10.7), Parkinson’s disease (N=50, Mean=63.4, SD=5.21), and osteoporosis (N=51, Mean=64.1, SD=7.25). Even the cohort with the lowest mean age among cases, chronic kidney disease (N=50, Mean=41.1, SD=15.1), was older on average than the controls. Our control group, derived from the VN1K project, consisted of 1,008 individuals with a mean age of 37.7 (standard deviation [SD] 3.59) and a median age of 37.0 (range: 32.0–49.0). This cohort was designed to achieve a nearly balanced sex distribution, with 497 males (49.3%) and 511 females (50.7%). These significant age differences between most case groups and the control group are consistent with the age-related nature of these conditions and highlight the critical need to adjust for age as a covariate in our subsequent PRS association analyses. Body mass index (BMI) measurements revealed no substantial differences between control and case cohorts, with all groups exhibiting values within the normal weight range. The control group demonstrated a mean BMI of 22.8 kg/m² (SD 2.62), which closely aligned with values observed across disease cohorts. BMI values across disease groups ranged from the lowest in gastric cancer patients (Mean=20.2 kg/m², SD=3.04) to the highest in Hyperlipidemia patients (associated with metabolic disorder) (Mean=23.7 kg/m², SD=2.86), yet all mean values remained within normal distribution, without a significant shift. This anthropometric homogeneity across cohorts suggests that BMI is unlikely to represent a significant confounding variable in our analyses.

**Table 1:** Summary characteristics of individuals across cohorts.

|  | VN1K<br>(N=1008) | Breast Cancer<br>(N=99) | Colorectal Cancer<br>(N=100) | Gastric Cancer<br>(N=50) | Parkinson Disease<br>(N=50) |
| --- | --- | --- | --- | --- | --- |
| **Age** |  |  |  |  |  |
| Mean (SD) | 37.7 (3.59) | 48.5 (9.16) | 56.8 (9.53) | 58.3 (9.31) | 63.4 (5.21) |
| Median [Min, Max] | 37.0 [32.0, 49.0] | 47.0 [34.0, 70.0] | 60.0 [36.0, 75.0] | 61.0 [35.0, 70.0] | 64.5 [42.0, 70.0] |
| **Sex** |  |  |  |  |  |
| Male | 497 (49.3%) | 0 (0%) | 68 (68.0%) | 39 (78.0%) | 15 (30.0%) |
| Female | 511 (50.7%) | 99 (100%) | 32 (32.0%) | 11 (22.0%) | 35 (70.0%) |
| **BMI** |  |  |  |  |  |
| Mean (SD) | 22.8 (2.62) | 22.5 (2.76) | 21.3 (2.51) | 20.2 (3.04) | 21.9 (2.70) |
| Median [Min, Max] | 22.7 [15.2, 34.3] | 22.2 [16.6, 31.3] | 21.3 [16.4, 28.3] | 19.8 [15.6, 28.3] | 21.6 [15.6, 29.8] |
| Missing | 0 (0%) | 40 (40.4%) | 13 (13.0%) | 6 (12.0%) | 0 (0%) |

**Table 1: Summary characteristics of individuals across cohorts.**
|  | Chronic Kidney<br>Disease<br>(N=50) | Coronary Artery<br>Disease<br>(N=50) | Hyperlipidemia<br>(N=50) | Osteoarthritis<br>(N=50) | Osteoporosis<br>(N=51) |
| --- | --- | --- | --- | --- | --- |
| **Age** |  |  |  |  |  |
| Mean (SD) | 41.1 (15.1) | 66.7 (10.7) | 46.6 (6.60) | 53.8 (10.5) | 64.1 (7.25) |
| Median [Min, Max] | 41.0 [18.0, 70.0] | 67.5 [44.0, 88.0] | 47.0 [35.0, 64.0] | 52.5 [34.0, 73.0] | 66.0 [46.0, 85.0] |
| **Sex** |  |  |  |  |  |
| Male | 28 (56.0%) | 31 (62.0%) | 18 (36.0%) | 11 (22.0%) | 3 (5.9%) |
| Female | 22 (44.0%) | 19 (38.0%) | 32 (64.0%) | 39 (78.0%) | 48 (94.1%) |
| **BMI** |  |  |  |  |  |
| Mean (SD) | 20.6 (2.58) | 22.9 (2.57) | 23.7 (2.86) | 22.7 (2.86) | 21.7 (2.46) |
| Median [Min, Max] | 20.2 [15.8, 26.8] | 22.9 [18.6, 29.7] | 23.1 [18.8, 32.1] | 22.5 [18.0, 30.6] | 21.3 [16.9, 28.9] |
| Missing | 1 (2.0%) | 0 (0%) | 0 (0%) | 0 (0%) | 0 (0%) |

### Establishing a widely applied workflow for PRS and TRS calculation and evaluation

We established a rigorous and comprehensive end-to-end workflow, consisting of 1) data curation including GWAS, LD references, genotype, and other external data, 2) data processing including merging, annotation and quality control, and 3) the execution of both fundamental and advanced PRS methodologies with diverse parameter settings. This robust workflow enabled an in-depth evaluation of model performance on our target dataset. For each method employed, we standardized the input from GWAS summary statistics to meet specific requirements. Posterior effect sizes were stored, and PRS were calculated using the PLINK tool ^28^. The resulting PRS were subsequently evaluated using liability R² and AUC metrics (**Figure 1**). The workflow is briefly described below as part of the results, with the focus on genotype data pre-processing, GWAS data curation, and PRS and TRS calculation.

**Fig 1:**
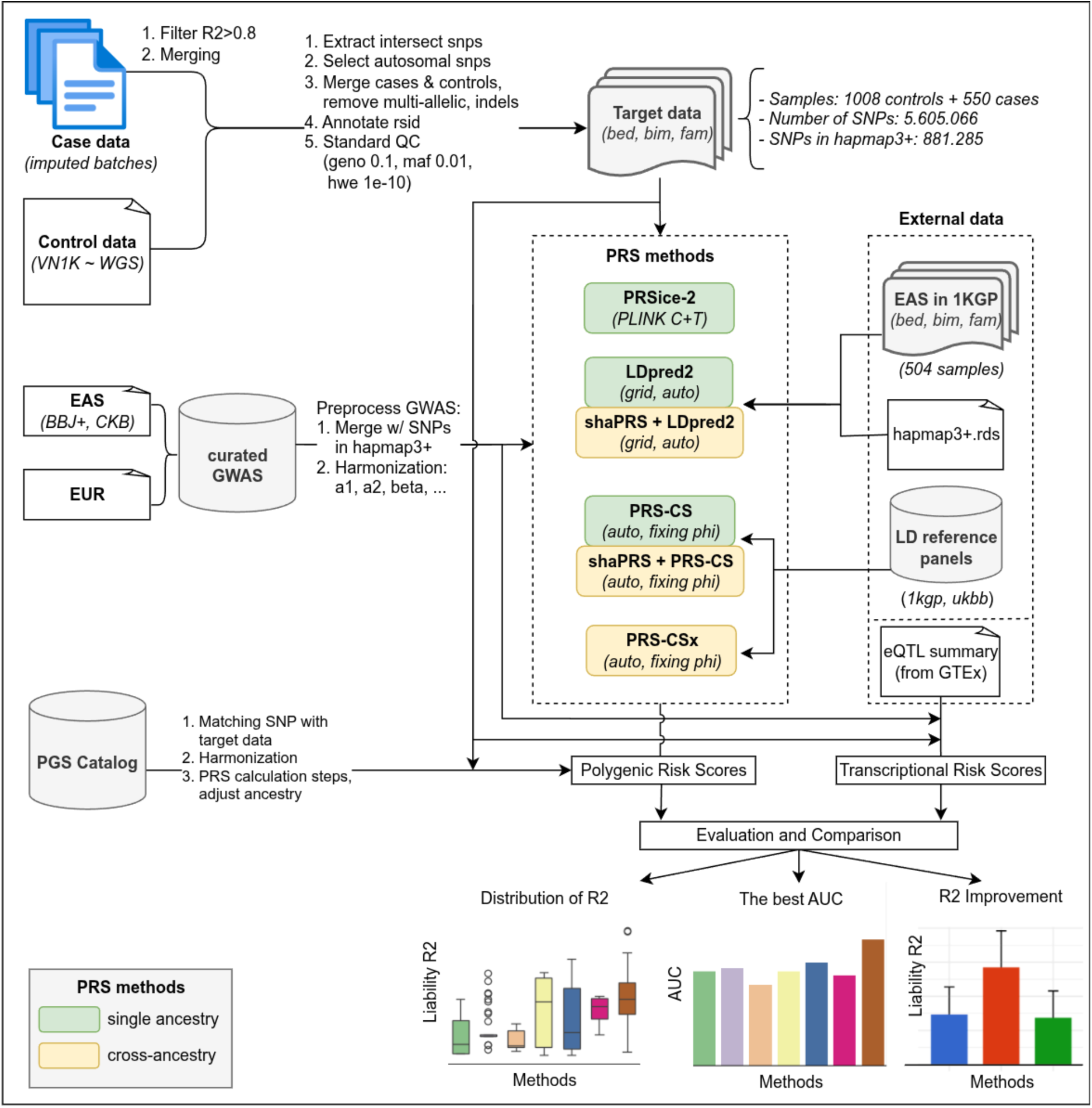
Comprehensive workflow for PRS and TRS calculation and evaluation. **Top**: Case data from nine diseases and control data were collected. After preprocessing steps, target data in Plink formats includes 1,558 samples and over 5.6 million SNPs, among which 881,285 SNPs overlap with Hapmap3+ ^29^. **Middle-Left**: Curated GWAS from EAS, EUR and PGS Catalog were used. **Middle-Right**: Multiple PRS methods including 3 single ancestry methods and 2 cross ancestry methods with different requirement files and parameters. In addition, TRS was integrating with data from GTEx. **Bottom-Right**: Evaluation and comparison were performed and resulted in distribution of R^2^. The best AUC and R^2^ improvement compared with the null model was reported.

#### Genotype data pre-processing

The patient genotype data was imputed in batches alongside other healthy individuals (with each batch containing 96 samples) using the 1KGP_VN1K reference panel, which combined 2,504 samples from the 1KGP project ^30^ and 1,008 samples from the VN1K project ^19^. We filtered out SNPs with R² ≤ 0.8 in each batch and then merged them together. This data was subsequently combined with the whole genome sequencing (WGS) data from 1,008 healthy individuals (VN1K). The quality control (QC) steps included extracting intersecting SNPs, selecting autosomal SNPs, merging cases and controls data, removing multi-allelic variants and indels, and annotating rsID (dbSNP 151) ^31^. Finally, we further filtered SNPs to include those with missing genotype rates < 10% (--geno 0.1), minor allele frequency (MAF) > 0.01 (--maf 0.01), and Hardy-Weinberg equilibrium p-values > 1e-10 (--hwe 1e-10) using the PLINK tool (v1.9). The post-QCed data was saved in PLINK file format (.bed, .bim, .fam) which included 1,558 samples (1,008 controls and 550 cases across nine diseases) and 5,605,066 SNPs. Among these, 881,285 SNPs were part of the HapMap3+ variants. We utilized this data as the target data for subsequent PRS calculations.

#### GWAS data curation

The raw materials for our PRS calculations consisted of GWAS from the East Asian populations. We utilized summary statistics referred to as BBJ+ that provided by BioBank Japan (for BC, CRC, GC, CAD, CKD and osteoporosis) and derived from East Asian populations available in GWAS Catalog for hyperlipidemia (Taiwanese), Parkinson disease (Korean) and osteoarthritis (detailed in **Supplement Table 1**). In addition, we curated Europe GWAS data (UKB) from GWAS Catalog and integrated them with summary statistics data in BBJ+ for a cross-ancestry approach (PRS-CSx, shaPRS).

#### PRS and TRS calculation

We used various common PRS methods, including three single ancestry methods (PRSice-2, LDpred2, PRS-CS), two cross-ancestry methods (shaPRS, PRS-CSx), and the combinations of them. The PRSice-2 method was the most simple and fast to run. This program was written in R language which implemented the standard C+T method, performs high-resolution scoring and generates the “*best-fit*” PRS following the difference of R² automatically. The LDpred2 and PRS-CS were Bayesian methods which computed posterior SNP effect sizes in HapMap3 ^32^ or HapMap3+ variants. We used LDpred2-auto and LDpred2-grid, which calculated LD correlation matrix from 504 individuals in East Asia of 1KGP - the same genomic ancestry as individuals in the GWAS, performed LD score regression and tune hyper-parameters for the validation data. PRS-CS is a command-line tool implemented in Python that estimates posterior SNP effect sizes using continuous shrinkage (CS) priors, based on GWAS summary statistics and an external linkage disequilibrium (LD) reference panel. PRS-CSx was a recent method developed from PRS-CS, that integrates GWAS and LD reference panels from multiple populations to improve cross-ancestry polygenic score prediction. The shaPRS method was used as a preprocessing GWAS step with proximal and adjunct ancestry to improve performance for downstream PRS methods (shaPRS + LDpred2 and shaPRS + PRS-CS). We developed a workflow to preprocess data, integrate summary statistics and methods to estimate SNP effect sizes and evaluate them with our genotypic data. Additionally, we utilized the *pgs-calc* software to curate PGS catalog sumstats from the PGS Catalog for the GRCh38 genome build, and to select summary statistics based on EFO_IDs for the nine diseases. Variants in the scoring files were then harmonized against those in the target dataset to account for strand orientation and allele matching, followed by the calculation of the PRS using the harmonized data. Besides evaluating PRS, we aimed to investigate how transcriptomic regulation contributes to disease risk by constructing a TRS. We used the best-performing PRS model from BioBank Japan (BBJ) GWAS summary statistics and integrated tissue-specific eQTL data from GTEx v8. To prioritize genes with putative causal links to disease, we performed colocalization analysis using the *coloc* R package ^13^. For each SNP-gene pair, we assigned regulatory direction (high or low expression) based on the effect of the risk allele then calculated TRS by summing polarized Z-scores across retained genes per individual. Finally, we re-evaluated TRS from selected tissues to the PRS-based model and assessed model performance using liability R² and AUC to quantify the incremental predictive value of transcriptome-informed genomic risk.

### Evaluating PRS performances with East Asian-centric GWAS data

The predictive capacity of polygenic risk scores (PRS) constructed using East Asian-centric GWAS data (BBJ+) was systematically evaluated across nine common diseases, with performance quantified by liability-scale pseudo-R² (**Figure 2**). These analyses, adjusted for sex and the first ten principal components (PC1-10) but excluding age to isolate the genomic contribution, revealed a spectrum of PRS performance both across diseases and among the seven PRS methodologies. The maximum phenotypic variance explained by any PRS model compared to the null model ranged from a modest 1.8% for chronic kidney disease (CKD) to a more substantial 8.3% for coronary artery disease (CAD).

**Fig 2:**
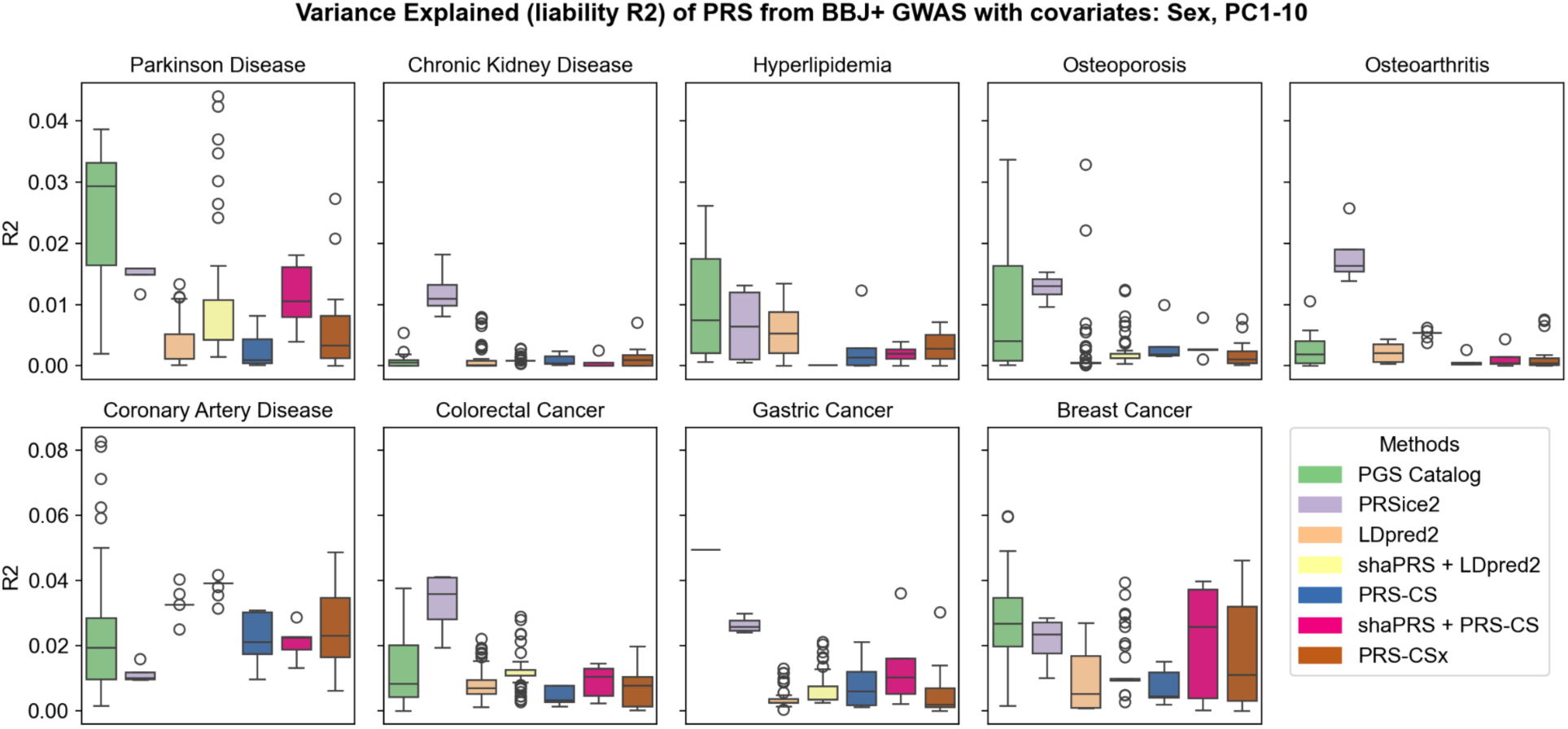
Variance explained (liability-scale pseudo-R²) of polygenic risk scores (PRS) using GWAS data from BioBank Japan and other East Asian sources (BBJ+), with sex and PC1-10 as covariates. The boxplots show the first to the third quartile of lncR^2^ for each method compared with the null model and the whiskers reflect the maximum and minimum values within 1.5 × interquartile range for each group.

PRSice-2, which implements a clumping and thresholding (C+T) approach, frequently yielded the highest R² values for several diseases, notably CKD (R^2^=0.018, 95%CI=0.002-0.034, P-value=0.025), osteoarthritis (R^2^=0.026, 95%CI=0.007-0.045, P-value=7.2×10^−3^), colorectal cancer (CRC) (R^2^=0.041, 95%CI=0.018-0.064, P-value=4.1×10^−4^), and breast cancer (BC) (R^2^=0.060, 95%CI=0.023-0.096, P-value=1.2×10^−3^). This marked improvement highlighted the importance of appropriate methods adapted to the genomic architecture of the diseases. C+T has shown a higher accuracy for traits with sparse genomic architecture. C+T may match well with traits where only top GWAS hits are informative in the target population.

In contrast, PRS derived from the PGS Catalog exhibited considerable variability in their R² distributions, particularly for Parkinson’s Disease (PD) (R²=0.039, 95%CI=0.016-0.061, P-value=8.5×10^−4^), hyperlipidemia (R²=0.026, 95%CI=0.007-0.045, P-value=6.7×10^−3^), CAD (R²=0.083, 95%CI=0.051-0.115, P-value=3.1×10^−7^). This wide dispersion reflected the heterogeneity of the curated scores, which originate from diverse ancestral backgrounds, discovery sample sizes, and analytical pipelines. While some cataloged scores demonstrated strong concordance with the Vietnamese target data, others, likely developed in populations with differing linkage disequilibrium structures or allele frequencies, showed attenuated predictive power. This observation underscored the critical importance of ancestry matching or the application of robust trans-ethnic methodologies when utilizing pre-computed PGS.

The Bayesian methods - LDpred2 (both *‘grid’* and *‘auto’* implementations), PRS-CS, and the cross-ancestry PRS-CSx along with their shaPRS enhanced counterparts (shaPRS+LDpred2 and shaPRS+PRS-CS), generally presented a more varied performance spectrum when considering the full range of their tested parameterizations, as visualized by the spread of their respective boxplots. These methods derived posterior SNP effect sizes independently of the target phenotype data, treating it as a true test set. Consequently, their R² distributions often encompassed a broader range of outcomes, indicating sensitivity to hyperparameter choices (e.g., the proportion of causal variants ‘*p*’ and heritability ‘*h²*’ in LDpred2-grid, or the global shrinkage parameter *‘phi’* in PRS-CS). While no single Bayesian approach consistently showed dominating performance, LDpred2 occasionally yielded competitive R² values, for instance in Osteoporosis (R²=0.033, 95%CI=0.012-0.054, P-value=2.2×10^−3^). The shaPRS meta-analytic framework was designed to improve trans-ethnic PRS portability by integrating EAS (proximal) and EUR (adjunct) GWAS summary statistics. It showed modest and disease-specific improvements when combined with LDpred2 or PRS-CS (as seen in PD, colorectal cancer, gastric cancer and breast cancer), suggesting its utility is contingent upon the specific genomic architectures and the degree of shared genomic effects between the ancestral groups. Notably, PRS-CSx did not demonstrate the expected superior performance compared to single-ancestry methods across most diseases, potentially due to the absence of a linear combination optimization step with training data, which is crucial for accurately calibrating ancestry-specific effect coefficients.

We benchmarked R² of polygenic risk scores (PRS) in various methods, including LDpred2, shaPRS + LDpred, PRS-CS, shaPRS + PRS-CS, PRS-CSx and PRSice2, using GWAS data from BBJ+. The covariates included sex and the first ten principal components (PC1-10). The variance explained (R²) was calculated by subtracting the R² of the null model (phenotype ∼ covariates) from the R² of the PRS model (phenotype ∼ covariates + PRS). Additionally, we computed the pseudo-R² metric, which accounts for the case/control ratio and is measured on the liability scale. The case and control groups differed significantly in mean age (as in **Table 1**), leading to a substantial contribution of the **’age’** covariate to the overall variance. To assess the independent genomic effect captured by the PRS, we excluded age from the covariates. For enhanced visualization, we organized the first eight diseases into the same range of y-axis values (due to their small R² values), while the subfigure of breast cancer used a different range (due to their larger R² values). The similar reports with covariates such as sex, age and PC1-10 are presented in **Supplementary Figure 1**.

For each disease, seven distinct algorithms were sequentially implemented to compute polygenic risk scores (PRS), with each method calculating scores based on its specific sets of parameters. Each parameter set yields a unique set of scores for the case and control cohorts, wherein every individual is assigned a corresponding score. The total number of such score sets is contingent upon the number of parameter configurations inherent to each method. Specifically, for the PGS Catalog, the quantity of score sets vary per disease, owing to differences in the data curation process for each trait within the catalog. The PRSice2 method generated four score sets, corresponding to clump-r^2^ values of 0.1, 0.4, 0.7, and 0.9. The LDpred2 method yields a maximum of 91 score sets, comprising one set from “LDpred2-auto” and up to 90 sets from “LDpred2-grid” (detailed parameter information is provided in the Methods section); in some instances, LDpred2-grid may fail to yield posterior beta estimates if the parameter combinations in the grid (e.g., heritability, h2, and p-value threshold, p) are incompatible with the trait’s genomic architecture or if convergence issues arise from a high h2 coupled with a low p-value. The PRS-CS method produced five score sets corresponding to the global shrinkage parameter (phi) values of 1e^-6^, 1e^-4^, 1e^-2^, 1, and “auto” setting. The number of scores for the shaPRS+LDpred2 and shaPRS + PRS-CS methods is analogous to that of LDpred2 and PRS-CS, respectively. For PRS-CSx, 15 score sets were generated, derived from the five phi parameters as used in PRS-CS, with each parameter applied to three sets of posterior effect sizes: specific to the East Asian (EAS), European (EUR) population, and combined across populations using an inverse-variance-weighted meta-analysis (META). Finally, a corresponding liability-scale R^2^ value was calculated for each of these generated score sets.

### Leveraging transcriptomic data to improve risk prediction accuracy

To capture tissue-specific effects on polygenic risk, we computed TRS separately for each disease using eQTL summary statistics from relevant tissues. Since eQTLs reflect how genomic variants regulate gene expression in specific tissues, using tissue-matched eQTL data allows TRS to prioritize SNP-gene pairs that are functionally active in the biological context relevant to the disease. This approach refines the interpretation of genomic risk by integrating not only the presence of risk alleles but also their regulatory impact on gene expression in disease-relevant tissues. Consequently, the resulting TRS provides a biologically informed, tissue-specific polygenic signal, complementing standard PRS and improving specificity in risk prediction.

Among the tissue-specific TRS evaluated for each disease, we identified the top-performing tissues whose inclusion yielded the greatest improvement in predictive accuracy. In the cancer group, breast cancer showed marked improvement when incorporating TRS from muscle tissue, with a 55% increase in AUC (R² = 0.071, 95% CI = 0.032–0.111, P = 3.54×10⁻⁴). For colorectal cancer, uterus-derived TRS resulted in a 33% AUC increase (R² = 0.043, 95% CI = 0.020–0.067, P = 2.93×10⁻⁴), while gastric cancer prediction improved by 69% using TRS from the stomach (R² = 0.056, 95% CI = 0.029–0.083, P = 4.40×10⁻⁵). Notably, coronary artery disease (CAD) prediction benefited most from TRS derived from the uterus, with a 49% improvement in AUC (R² = 0.120, 95% CI = 0.083–0.157, P = 1.40×10⁻¹⁰), highlighting a non-obvious regulatory tissue association. Chronic kidney disease (CKD) showed a modest improvement (11%) using TRS from the testis (R² = 0.019, 95% CI = 0.002–0.035, P = 2.31×10⁻²). Among neurological and age-related conditions, Parkinson’s disease prediction improved by 20% with TRS from the cerebellar hemisphere (R² = 0.062, 95% CI = 0.034–0.091, P = 1.42×10⁻⁵), consistent with the known role of this brain region in motor control. Osteoporosis and osteoarthritis were best predicted using TRS from liver (21%, R² = 0.036, 95% CI = 0.014-0.058, P = 1.39×10⁻³) and brain cerebellum (64%, R² = 0.033, 95% CI = 0.012-0.054, P = 2.12×10⁻³), respectively. Detailed improvements in AUC and R² across a broader set of tissue-disease pairs are provided in **Supplementary Table 3**, while the results for the top tissue associated with each disease are summarized in **Supplementary Table 4**.

Building on the tissue selection criteria described in the Methods, we constructed a joint PRS+TRS model by integrating the best-performing PRS with TRS derived from all selected tissues for each disease. We then compared the predictive performance of three models (1) PRS vs. Null, (2) PRS+TRS vs. Null, and (3) PRS+TRS vs. PRS across nine diseases. The joint PRS+TRS model consistently demonstrated higher liability R² than the PRS-only model, with improvements ranging from 0.01 to approximately 0.15 (**Figure 3**). The extent of improvement varied by disease, influenced by both the baseline predictive power of PRS and the contribution of TRS from different tissues (**Supplementary Table 3**). The largest improvements were observed for CAD (R² = 0.151, 95% CI = 0.111-0.191, P = 8.484.40×10^-14^), hyperlipidemia (R² = 0.161, 95% CI = 0.121-0.202, P = 5.39×10^-15^), gastric cancer (R² = 0.144, 95% CI = 0.105-0.184, P = 4.18×10^-13^), breast cancer (R² = 0.108, 95% CI = 0.061-0.154, P = 4.86×10^-6^), and osteoporosis (R² = 0.0065, 95% CI = 0.037-0.094, P = 7.88×10^-6^), reflecting a substantial contribution of TRS to predictive power. Conversely, CKD (R² = 0.001, 95% CI = 0-0.003, P = 0.704), osteoarthritis (R² = 0.012, 95% CI = 0-0.025, P = 0.071), and Parkinson’s disease (R² = 0.041, 95% CI = 0.018-0.065, P = 0.0005) showed smaller gains, potentially due to fewer contributing tissues in the TRS model.

**Fig 3:**
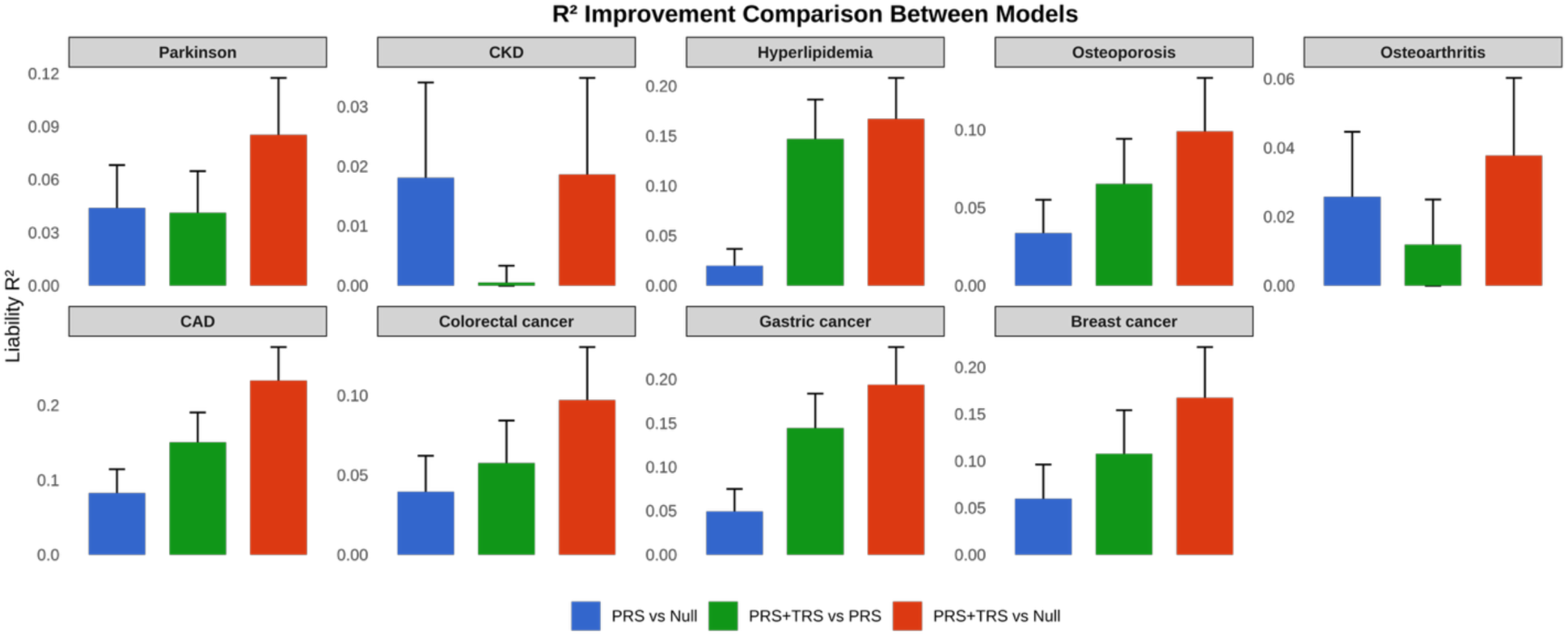
Comparison of variance explained (liability R²) between PRS+TRS, PRS, and Null models with covariates sex and PC1-10 for nine diseases. The Null model includes only covariates (sex and PC1–10; for breast cancer, only PC1–10). The PRS model adds the best PRS from BBJ+, while the PRS+TRS model further incorporates the TRS from selected tissues. Liability R² differences were computed by directly subtracting the liability R² values between the corresponding models. Bar colors indicate model comparisons: blue for PRS vs. Null, green for PRS+TRS vs. PRS, and red for PRS+TRS vs. Null. Error bars denote the 95% confidence intervals. These results demonstrate the additional variance explained by integrating TRS with PRS, beyond the contributions of PRS or covariates alone.

The overall variance explained by the models was higher than typically reported in previous studies ^9, 33^. This may be influenced by the exclusion of age as a covariate, which is known to impact all nine diseases. The extent of improvement varied across diseases, reflecting differences in PRS performance and the relevance of TRS derived from multiple GTEx tissues.

### Implications on clinical impact and risk stratification results across the nine diseases

PRS offers a promising tool to stratify individuals by their inherited risk for complex diseases, enabling early identification of high-risk groups who may benefit from targeted screening or preventive strategies. Previous studies have shown that the predictive power of PRS varies across age and sex ^9, 34, 35^, emphasizing the value of integrating clinical covariates into risk models. In our study, we investigated the potential clinical utility of PRS-based risk stratification across nine common diseases in the Vietnamese population. By categorizing individuals into PRS percentiles, we observed a clear and progressive increase in disease odds ratios for those in higher risk strata compared to the bottom 20% reference group. This pattern held across most diseases tested and was particularly evident in certain subgroups, such as males, where the risk differential was even more pronounced. Such stratification suggests potential clinical implications: individuals in the top PRS decile may warrant enhanced surveillance, lifestyle counseling, or earlier diagnostic testing, even before traditional clinical risk factors become apparent. For example, women with high breast cancer PRS could benefit from earlier mammography screening, while those at high genomic risk for CAD may be considered for more aggressive lipid control. Furthermore, PRS may help refine risk prediction when combined with traditional factors in clinical decision-making algorithms. As PRS methodologies continue to evolve, integrating age, sex, and ancestry-specific data will be crucial to maximizing their clinical relevance. Our findings demonstrate the feasibility and potential value of PRS in guiding personalized preventive care and resource allocation in a population that has been historically underrepresented in genomic studies.

We next aimed to assess how the predictive power of PRSs for these nine diseases may vary by sex and age groups, thereby evaluating the advantages of genomic risk profiling that takes these clinical characteristics into account. In **Figure 4**, for each disease, we first categorized individuals based on their PRS values into distinct percentile groups. This adaptation to wider percentile bins for the sex-stratified analysis was implemented to ensure robust estimation by maintaining adequate sample sizes within each sub-category, thereby mitigating the risk of deriving unstable odds ratios from sparsely populated groups.

**Fig 4:**
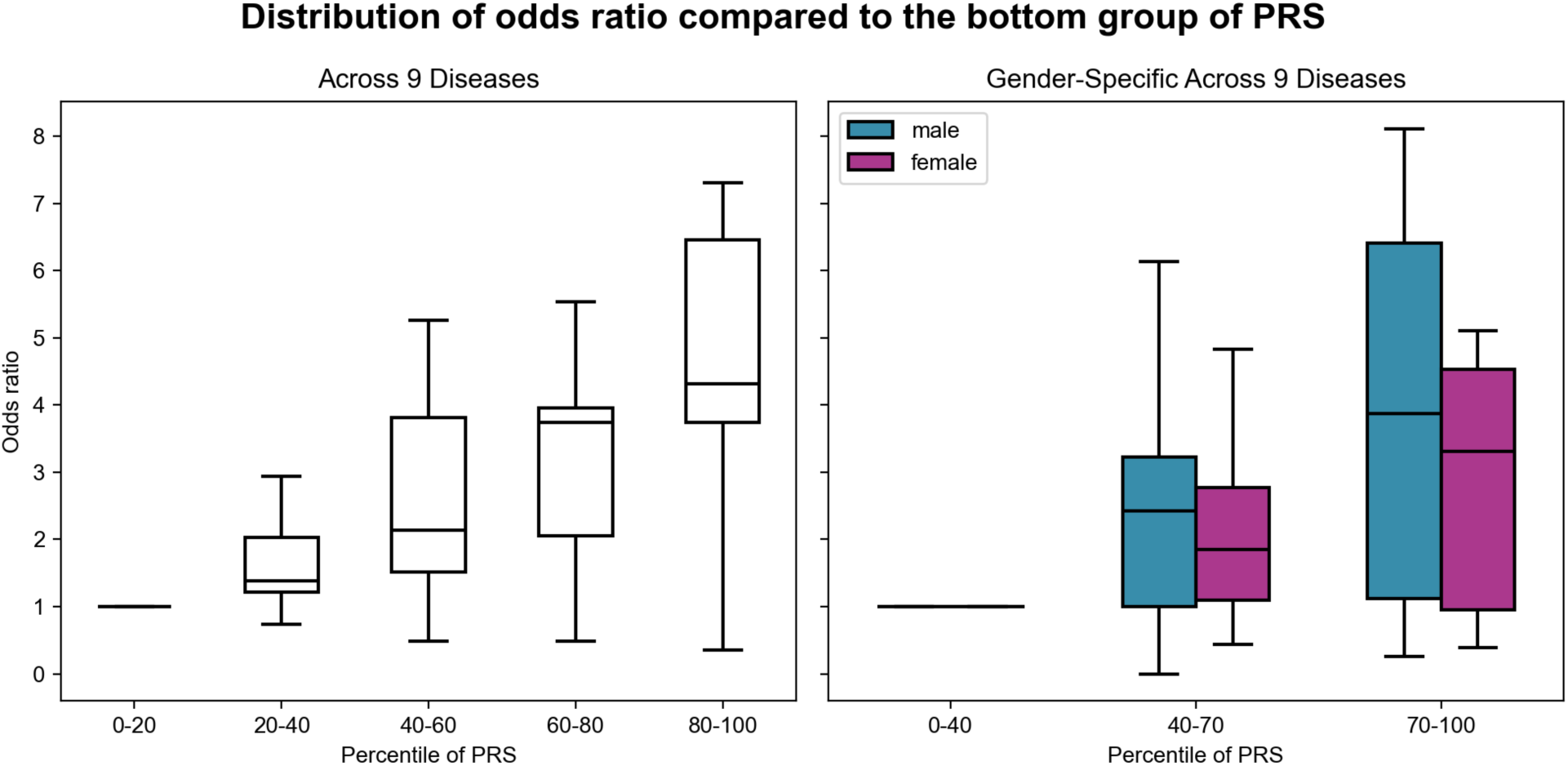
Distribution of odds ratio compared to the bottom group of PRS. The right panel presents these odds ratios for PRS quintiles across 9 diseases, specifically: 0-20th (baseline), 20th-40th, 40th-60th, 60th-80th, and 80th-100th percentiles. The left panel shows gender-specific stratum, where the cohort of 1,558 individuals were categorized into three broader PRS percentile groups: 0-40th (baseline), 40th-70th, and 70th-100th. The boxplots show the first to the third quartile of odds ratio for nine diseases and the whiskers reflect the maximum and minimum values within 1.5 × interquartile range for each group.

**Figure 4** illustrates the clinical potential of PRS for risk stratification across nine diseases studied in the Vietnamese population. Overall, within each PRS quintile, the differences in odds ratios between diseases are relatively substantial, as demonstrated by the wide range of values across the boxplots. The first panel demonstrates a consistent dose-response relationship: as individuals fall into increasingly higher PRS quantiles, their disease prevalence increases significantly. Specifically, individuals with genomic risk in the highest quintile (80th-100th percentile) exhibit an average disease risk more than 4-fold higher compared to those in the lowest quintile (0-20th percentile). This marked disparity emphasizes the capability of PRS to effectively stratify populations into distinct risk categories. Further, the second panel provides assessments of increased disease risk across PRS percentile groups stratified by sex. While both males and females exhibit a similar pattern of increasing odds ratios with higher PRS strata, the data suggests potentially steeper risk gradients for males. Men in the top PRS group (70-100th percentile) exhibit higher odds ratios than their female counterparts within the same genomic stratum. This observation, although requiring further investigation with larger sample sizes to definitively confirm differential effects, suggests that the impact of polygenic risk on disease manifestation may quantitatively differ between sexes.

Collectively, the stratification patterns depicted in **Figure 4** demonstrate that the Vietnamese population exhibits distinct predispositions to common diseases. Disease incidence escalates with increasing genomic risk, a gradient potentially more pronounced in males.

## DISCUSSIONS

In this study, we leveraged several advanced PRS methods to predict susceptibility to nine common diseases within the Vietnamese population. Given the historical underrepresentation of Southeast Asian populations in genomic research ^36, 37^, our study provided critical insights into the development and potential application of PRS in such under-studied populations. We also demonstrated that integrating PRS with TRS provided added predictive value on the Vietnamese population, therefore benefit underrepresented populations.

The variance explained (R²) by PRS models in this study ranged from 1.8% to 8.3%, indicating that PRS can effectively identify individuals with a high genomic risk. These findings are consistent with previous studies that demonstrated reduced PRS transferability from European-derived models to non-European populations, reflecting differences in linkage disequilibrium patterns and allele frequencies ^38, 39, 40^. The AUC values ranged between 0.55 (for CKD) to 0.70 (for CAD), suggesting that PRS might be considered a useful tool for risk stratification, particularly for identifying individuals who may benefit from targeted interventions. Among the PRS methods evaluated, PRSice-2 showed consistently better performance across certain diseases, including colorectal cancer, chronic kidney disease, osteoarthritis, and breast cancer. This result likely stems from the method’s ability to optimize SNP selection dynamically through clumping and thresholding procedures tailored to the target data. In contrast, Bayesian approaches such as LDpred2 and PRS-CS, even though considered to be more theoretically sophisticated, exhibited greater variability across diseases and parameter settings. This variability highlighted the necessity of careful selection of method and optimization of parameters, especially when applying PRS methodologies to populations with different genomic ancestries.

We also assessed the performance of cross-ancestry PRS methods (PRS-CSx, shaPRS), which integrated GWAS data from both East Asian and European populations. However, we found that these methods did not consistently enhance predictive performance across all diseases studied. This finding suggests persistent complexities arising from ancestral differences, such as variation in linkage disequilibrium patterns and allele frequencies, which limited the effectiveness of the current cross-ancestry approaches. Future developments in PRS methodologies specifically tailored for Southeast Asian populations are thus warranted to help overcome such limitations and enhance predictive performance.

Furthermore, our findings also demonstrated potential clinical utility of PRS for risk stratification. Individuals with higher PRS values exhibited significantly increased their odds ratios for having such diseases. This trend was further stratified by sex, emphasizing the relevance of integrating clinical characteristics with genomic risk information in personalized medicine. The capacity to discern individuals at substantially elevated risk even prior to clinical manifestation based on their genomic profiles holds a considerable potential for advancing personalized and preventive healthcare. In principle, such genomic risk information could inform tailored screening schedules, guide lifestyle modification counselling, or prioritize individuals for early intervention strategies. This evidence strongly supports the integration of PRS into broader clinical risk assessment frameworks, paving the way for more personalized approaches to disease prevention within the Vietnamese population.

Our study also demonstrated the added predictive value of TRS on an underrepresented population. By integrating PRS with tissue-specific TRS derived from the most relevant tissues for each disease, we consistently observed improved predictive accuracy across all nine diseases. Notable examples include a 15% increase in liability R² for coronary artery disease and hyperlipidemia, as well as substantial improvements for gastric cancer (14%) and breast cancer (10%). These findings indicate that TRS despite being constructed from eQTL data not specific to the Vietnamese population can complement PRS and enhance complex disease prediction, even within the constraints of a modest sample size.

This study has several limitations. First, interpretation of our data should be cautious as we had a relatively small sample size, limiting statistical power and generalizability. Future studies on population-specific genomic resources and conducting larger validation studies within the Vietnamese population are therefore warranted to improve the robustness and accuracy of PRS-based predictions. Second, we observed significant differences in the age distributions between case and control groups for several diseases. This age discrepancy, if not properly accounted for, might potentially influence the interpretation of PRS performance, since age itself is a strong risk factor for such diseases. Third, the TRS used in our study were constructed using eQTL summary statistics derived from European samples, which may not accurately reflect gene regulation patterns specific to the Vietnamese population. And we also lacked RNA-sequencing data to directly derive TRS from our cohort, which could have enabled more accurate modeling of transcriptomic contributions to disease risk. Fourth, PRS/TRS hold potential for risk stratification, disease prediction, and personalized prevention in the Vietnamese population, but they should be considered as complementary tools rather than replacements for traditional clinical risk factors. Large-scale studies integrating genomic, clinical, and environmental factors are needed to assess their true clinical impact before widespread implementation.

In conclusion, our study highlights potential applications of PRS and TRS models for disease risk prediction in the Vietnamese population, which is underrepresented in global genomic database. Future research should prioritize large-scale, ancestry-specific genomic datasets, refined cross-ancestry methodologies, and multi-omics integrations such as TRS. These advancements would significantly enhance personalized risk assessment and healthcare outcomes in diverse populations, especially populations residing in the Southeast Asian region.

## METHODS

### Sample collection

Sample collection was conducted at the clinical departments of four hospitals in Hanoi, Vietnam. A total of 550 patients with nine chronic diseases were screened by specialists and physicians, assisted by nurses, and those who met the criteria and provided consent signed an informed consent form (ICF). Clinical staff collected patient information using CRF forms provided by the research team, and only patients with complete data were included. Blood samples with 2ml volume were collected in a single EDTA tube and stored at 2-4°C. Within 24 hours of receiving the documents from the hospitals, research assistants of the project entered ICF data into the RedCap system hosted by Hanoi Medical University (https://khaosat.spmph.edu.vn/redcap/). Each electronic medical record (EMR) was tailored to the specific disease and facility, including patient demographics, diagnostic and exclusion criteria, medical and family history, risk factors, treatment details, diagnostic tests, and adverse event monitoring. Samples were collected from Bach Mai Hospital for coronary artery disease, chronic kidney disease, and dyslipidemia; the National Geriatric Hospital for Parkinson’s disease, osteoporosis, and osteoarthritis; and Hanoi Medical University Hospital and Viet-Duc Hospital for breast cancer, colorectal cancer, and stomach cancer. This study was approved by the Institutional Review Board of the Hanoi Medical University, Hanoi, Vietnam - IRB-VN01.001/IRB00003121/FWA00004148 (*Approval No.: 362/GCN-HĐĐĐNCYSH-ĐHYHN*)

### SNP genotyping of disease cohorts

Genomic DNA was extracted from peripheral blood samples collected from study participants using QIAamp DNA Blood Mini Kit (Qiagen, Dusseldorf, Germany) following the described protocol ^41^.DNA quantification was carried out using either the Qubit dsDNA BR Assay Kit (Thermo Fisher Scientific, Waltham, USA) as previously ^42^, ensuring total DNA amount exceeded 1 μg and the concentration of DNA was above 20 ng/μL. Genotyping was performed using the Axiom Precision Medicine Diversity Research Array following the standard Axiom 2.0 workflow on the Applied Biosystems™ GeneTitan™ Multi-Channel Instrument. Each batch comprised 96 samples, with blinded and positive control samples randomly incorporated to assess data consistency. Genotyping accuracy was confirmed, with all control samples showing concordance rates above 98%. Raw data files (CEL format) were processed using the Applied Biosystems™ Axiom™ Analysis Suite software, following the guidelines in the Axiom Genotyping Solution Data Analysis Guide. The generated genotype calls were converted into VCF format for further analysis. To enhance variant resolution, genotype imputation was performed using a reference panel built from 1,008 whole-genome sequences in the VN1K dataset, retaining only high-confidence imputed variants with an R² threshold of ≥ 0.8. The final dataset was constructed by integrating 1,008 control samples from the VN1K project with 500 case samples, following the same processing and quality control steps outlined above. This combined dataset served as the target cohort for subsequent analyses.

### PGS Catalog data curation

We curated PGS summary statistics for our nine diseases from the PGS Catalog, focusing on entries for the GRCh38 genome build and selecting them based on the EFO trait IDs listed in **Supplementary Table 2**. The entire workflow, encompassing scoring file curation, variant matching, allele harmonization, and PGS calculation, was conducted using the pgsc_calc tool. First, variants in the scoring files were matched to those in the target datasets to ensure alignment between base and target data. Next, a harmonization step was performed to address strand orientation and allele matching issues, ensuring consistency across datasets. Finally, polygenic scores were computed by summing the effect sizes of the risk alleles identified in the harmonized data. To account for genomic ancestry, we used reference panels from the 1000 Genomes Project (1KGP) and 1KGP_HGDP to derive genomic ancestry data through principal component analysis (PCA). For each target sample, the most similar population in the reference panel was identified based on PCA results. Polygenic scores were adjusted for genomic ancestry using these population assignments to correct for population stratification effects.

### Polygenic risk score calculation

#### PRSice-2

this tool utilizes the standard C+T (Clumping and Thresholding) method which efficiently handles large-scale genome-wide association study (GWAS) summary statistics and enhances the precision of genomic risk estimation by addressing issues of linkage disequilibrium and significance thresholds. The PRS for an individual is calculated using the formula:

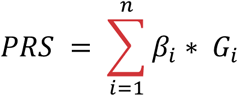

Where:

- *β_i_* is the effect size for SNP *i*.
- *G_i_* is the genotype for SNP *i* (usually coded as 0, 1, or 2, representing the number of risk alleles)

The clumping process in the PRSice-2 method utilizes a correlation measure known as *r*^2^ to assess the linkage disequilibrium (LD) between SNPs. We adjust the *’clump-r2’* parameter to values of 0.1, 0.4, 0.7, and 0.9 to evaluate the output results from PRSice-2. After clumping, SNPs were selected according to thresholds from 5e^-8^ to 1 with interval 5e^-5^ and were multiplied by genotype like formula (1). For each *clump-r2* parameter value, we extracted the best-fit PRS to compare with other methods.

#### LDpred2-auto and LDpred2-grid

these methods employ a Bayesian framework to estimate SNP effect sizes and adjust for local LD patterns. By utilizing variants from the HapMap3+ dataset (1,444,196 variants), these two methods incorporate a comprehensive reference panel that captures the genomic architecture of diverse populations which is crucial for accounting for correlations between SNPs. We used the genotypes of 504 individuals from East Asian (EAS) superpopulation in 1000 Genomes Project (1KGP) data to compute the LD correlation matrix.

Two these extensions of the LDpred model assume the following model for effect sizes:

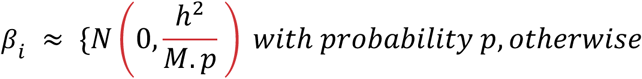

where *β_i_* is the effect size for SNP *i*, p is the proportion of causal variants, M the number of variants, ℎ^2^ is the heritability. LDpred2-auto estimates p and ℎ^2^ hyper-parameters within the Gibbs sampler. In total, we run LDpred2-auto 30 times with ℎ^2^*_ldsc_*, where ℎ^2^*_ldsc_* is the heritability estimate from the constrained LD score regression ^43^ as initial value for ℎ^2^ and a sequence of 30 values from 1e^-5^ to 0.5 equally spaced on a log-scale as initial values for p. In addition, we also use two new parameters for improving its robustness: *allow_jump_sign=FALSE* and *shrink_corr=0.95*. In contrast, LDpred2-grid uses several values of p and ℎ^2^ from a grid of hyper-parameters. We test a grid of values for p from a sequence of 15 values from 1e^-5^ to 0.5 on a log-scale, ℎ^2^ within {0.7, 1. 1.4}.ℎ^2^*_ldsc_* and whether sparsity is enabled or not.

#### PRS-CS and PRS-CSx

PRS-CS is a Bayesian method for polygenic risk prediction that estimates posterior SNP effect sizes from summary statistics through the use of a continuous shrinkage prior. This approach is effective across diverse genomic architectures, provides accurate modeling of linkage disequilibrium (LD), and is computationally efficient. This method involves a single hyperparameter, known as the global shrinkage parameter (*ϕ*), which represents the overall sparsity of the genomic architecture. In our analysis, we adopted the default parameter settings along with a pre-calculated LD reference panel that derived from HapMap3 variants with minor allele frequency (MAF) greater than 0.01, using EAS superpopulation samples from phase 3 of the 1KGP that matched with the ancestry of our discovery samples. We evaluated the global shrinkage parameter across values of 1e^-6^, 1e^-4^, 1e^-2^ and 1.0, selecting the value that maximized *R*^2^ as the performance metric for our phenotype validation dataset.

PRS-CSx is an extension of PRS-CS that allows for the integration of GWAS from multiple populations, enhancing polygenic prediction across different ancestries. The posterior effect size of SNP *j* in the population *k* is represented as a global-local scale mixture of normal distributions:

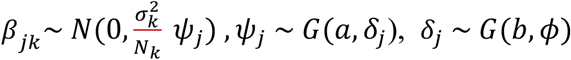

where *ϕ* is the global shrinkage parameter, *ψ_j_* is is the local shrinkage parameter for marginal associations of SNP *j*, *σ_k_* is the variance of non-genomic effects and *N_k_* is the number of individuals of population *k*; *G*(*α*, *β*) denotes gamma distribution with shape parameter *α* and scale parameter *β*, the default values in this work are set to a = 1 and b = 1/2. We run PRS-CSx by using GWAS and LD reference panels from BBJ+ (EAS) and EUR populations. We used with the global shrinkage parameter among 1e^-6^, 1e^-4^, 1e^-2^, and 1.0, along with the *META_FLAG* parameter is *True* to return combined SNP effect sizes across populations using an inverse-variance-weighted meta-analysis of the population-specific posterior effect size estimates. All individual-level polygenic scores can be calculated by concatenating output files from 22 chromosomes, which were evaluated by the maximum *R*^2^ in our phenotype validation dataset. Due to sample size limitations, we only performed stage 1 of PRS-CSx without optimizing using linear combination.

#### ShaPRS

this method offers a shrinkage-based approach to refine SNP effect sizes before downstream polygenic risk score (PRS) calculations, such as those performed by LDpred2 and PRS-CS. By integrating summary statistics from GWAS with reference panels that include both the proximal population (e.g., EAS) and adjunct population (e.g., EUR), shaPRS effectively adjusts for population-specific linkage disequilibrium (LD) patterns and allele frequency differences. This dual-population approach allows shaPRS to leverage the strengths of each reference group, reducing noise and overfitting in the effect size estimates, leading to more accurate and robust polygenic risk scores. Formula of final shaPRS SNP effect estimate as:

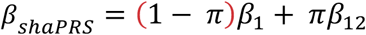

where *β*_12_ is the standard inverse variance weighted average of the effect size estimates in the proximal study (*β*_1_) and the adjunct study (*β*_2_), *π* is local false discovery rate (lFDR) as an estimate of the probability that effects are heterogeneous. We used the shaPRS method as a preprocessing GWAS step with proximal GWAS from BBJ+ (EAS) and adjunct GWAS from EUR (the same as PRS-CSx). This will output the final summary statistics file that we use as input for LDpred2 (shaPRS + LDpred2) and PRS-CS (shaPRS + PRS-CS).

### Colocalization Analysis using eQTL and GWAS Data

To prioritize genes whose expression may be causally linked to disease risk, we performed colocalization analysis using the coloc R package (version 5.2.3) in R 4.2.0. This analysis integrated genome-wide association study (GWAS) summary statistics from the BioBank Japan (BBJ) with tissue-specific expression quantitative trait loci (eQTL) summary statistics from the Genotype-Tissue Expression (GTEx) project (v8). The eQTL data was obtained from the eQTL Catalogue ^44, 45^, which provides harmonized QTLs across multiple tissues. Detailed information about the eQTL summary statistics, including sample size and tissue source, is provided in **Supplementary Table 2**. In our analysis, we included only eQTL datasets where the quantification method was based on gene expression. For each disease, we used the BBJ GWAS summary statistics corresponding to the best-performing polygenic risk score (PRS) model identified in our prior PRS evaluation step. Specifically, for each disease phenotype, we selected the PRS model with the highest predictive performance and then extracted the corresponding GWAS summary statistics to serve as input for colocalization.

For each gene, we matched SNPs across the GWAS and eQTL datasets to create two input datasets compatible with the *coloc.abf* function. The GWAS dataset contained p-values, effect sizes, standard errors, sample sizes, and MAF, and was modeled as case-control data (type = “cc”), with disease prevalence specified according to BBJ estimates. The eQTL dataset was modeled as a quantitative trait (type = “quant”), and included SNP-level p-values for gene expression associations, per-tissue sample sizes, estimated effect sizes, squared standard errors, and minor allele frequency. We performed Bayesian colocalization analysis with default parameters including p_1_=p_2_=1e^-4^ representing the prior probability that a SNP is associated with trait 1 and trait 2, respectively, and p_12_=1e^-5^ representing the prior probability that a SNP is associated with both traits.

Prior to colocalization, we filtered SNPs to retain only those with p-values < 1×10⁻⁴ in either the GWAS or eQTL dataset. When duplicate SNPs were present, we retained only the one with the most significant p-value. GTEx molecular trait IDs were mapped to gene symbols using the org.Hs.eg.db annotation package (version 3.16.0) ^46^.

We ran colocalization separately for each of the nine disease phenotypes. For each SNP-gene pair, the *coloc.abf* function computed the posterior probabilities of five hypotheses (*H*_3_ to *H*_4_). This method is based on the Bayesian test for colocalization ^13^, which evaluates whether two association signals are likely due to a shared causal variant. For each SNP, an Approximate Bayes Factor (ABF) is computed to quantify the evidence that the SNP is associated with a trait following the equation:

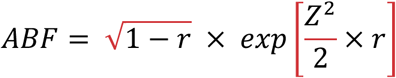

Where *Z* = 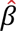/*SE* is the Z statistic for SNP-trait association, and the shrinkage factor 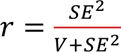 reflects the proportion of prior variance within the total variance from the assumption of a normal distribution *N(0, SE*^2^) on the effect size 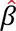. The prior variance *V* can be estimated from minor allele frequency and sample size from the summary statistics. Once the ABFs are computed for all SNPs and traits, they are used to calculate the posterior probabilities of five hypotheses:

- *H*_0_: Neither trait is associated
- *H*_1_: Only the first trait is associated
- *H*_2_: Only the second trait is associated
- *H*_3_: Both traits are associated, but with different causal variants
- *H*_4_: Both traits are associated and share the same causal variant The posterior probabilities are calculated using Bayes’ theorem:

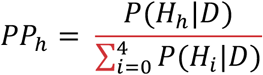

Where *PP_h_* is the posterior probability of hypothesis *h*, *D* is observed data from summary statistics.

We focused on hypothesis 4 (*H*_4_), which represents the scenario in which a single shared causal variant affects both gene expression and disease risk. A high posterior probability for *H*_4_ (≥ 0.8) indicates that the same variant likely underlies both the GWAS and eQTL associations, providing evidence for a causal regulatory mechanism. SNP-gene pairs with *H*_4_≥ 0.8 were retained as colocalized signals for downstream analysis.

### Transcriptional risk score calculation

Following colocalization, we removed any remaining duplicate SNP-gene associations by selecting the one with the most significant p-value. We then computed Z-scores from effect sizes (β) and standard errors (SE) according to the method described ^47^ where Z = β / SE.

To determine the direction of disease-associated gene regulation, we classified each gene as either “high expression” or “low expression” based on whether the disease risk allele was associated with increased or decreased gene expression, respectively. In rare cases where a gene was associated with SNPs in both directions (i.e., both high and low expression), the gene was excluded from downstream analysis to avoid conflicting effects, following the approach described ^15^.

Z-scores were standardized across genes to ensure a mean of 0 and variance of 1. For genes labeled as “*low expression*” we flipped the sign of their Z-scores, effectively aligning all gene expression changes with increased disease risk. This polarization allowed for additive modeling of gene-level effects on risk, regardless of the direction of regulation. The final TRS was computed for each individual by summing the polarized Z-scores across all retained SNP-gene pairs from the individual’s genotype data. TRS values were calculated separately for each disease using corresponding tissue-specific eQTL summary statistics.

### Tissue selection for TRS computation and PRS-enhanced model with TRS integration

To evaluate the contribution of tissue-specific TRS to disease risk prediction, we systematically assessed the improvement in model performance when integrating TRS with PRS across various tissues. Each model was adjusted for sex and top ten principal components (PC1-PC10); for breast cancer, only PC1–10 was included. The difference in predictive performance was quantified using AUC was measured as the difference between the AUC of the full model (PRS+TRS+covariates) and that of the null model (covariates only). To further quantify the relative gain in predictive power, we computed the proportional AUC improvement using the following equation:

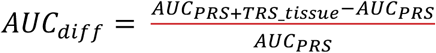

Tissues that yielded an increase in AUC relative to the optimal PRS baseline from the BBJ dataset were designated as selected tissues, denoting their consistent enhancement of model performance in the integrated PRS+TRS model. TRS from all selected tissues were incorporated into the final joint PRS+TRS model. We then evaluated prediction performance using liability R² values to compare three models, including 1) PRS vs. Null, 2) PRS+TRS vs. Null, and 3) PRS+TRS vs. PRS with the ultimate goal of quantifying the additional variance explained by integrating TRS into the PRS model. The Null model consisted of covariates including sex and the top ten genomic principal components (excluding sex for breast cancer). The PRS model extended the Null model by adding the PRS for each disease, while the PRS+TRS model further included TRS from all selected tissues.

## Data Availability

All data and results could be publicly found in https://github.com/SangGold97/VGP, and also the Main Text, Supplementary Files. Please contact the corresponding authors for further request.

## Competing interests

SVN, TMP, TTHT, GMV, NSV are current employees of GeneStory JSC, Vietnam, a company that develops and markets products for genetic testing. The other authors declare no competing interests.

## Author contributions

Initials of authors in the list (the same order): SVN, TMP, THH, TTHT, GMV, MHT, TKN, HLT, HTTV, TMP, DTN, AGP, YH, GHP, DXD, HNL, THT, QN, BT, NSV. NSV, QN, THT, and BT conceptualized, designed, and supervised the project. SVN, TMP, and THH conducted the primary data analysis and interpretation. TTHT and GMV contributed to the data analysis. MHT, THH, TKN, GHP, and DXD performed the data curation. MHT, THH, HLT, HTTV, TMP, DTN, AGP, YH, GHP, DXD, and TT contributed to the sample collection. MHT conducted the wetlab experiments. SVN, TMP, and THH drafted the manuscript. NSV, BT, QN, and HNL revised and wrapped the final manuscript. All authors read and approved the final manuscript.

## Acknowledgements

Our sincere appreciation goes to all the individuals and their families who generously agreed to participate in this research. We are grateful to the Hanoi Medical University Hospital, Bach Mai Hospital, Viet-Duc Hospital, National Geriatric Hospital, and National Heart Institute for their support with sample collection and clinical expertise. We also thank lab technicians at GeneStory who contributed to the genotyping experiments. This project was supported by VINIF Grant VINIF.DA.2020.02.

## Notes

### Competing Interest Statement

Sang V. Nguyen, Tien M. Pham, Trang T.H. Tran, Giang M. Vu, Nam S. Vo are current employees of GeneStory JSC, Vietnam, a company that develops and markets products for genetic testing. The other authors declare no competing interests.

### Author Declarations

This study was approved by the Institutional Review Board of the Hanoi Medical University, Hanoi, Vietnam - IRB-VN01.001/IRB00003121/FWA00004148 (Approval No.: 362/GCN-HĐĐĐNCYSH-ĐHYHN)

